# 3-generation family histories of mental, neurologic, cardiometabolic, birth defect, asthma, allergy, and autoimmune conditions associated with autism: an open-source catalogue of findings

**DOI:** 10.1101/2023.11.03.23298042

**Authors:** Diana Schendel, Linda Ejlskov, Morten Overgaard, Zeal Jinwala, Viktor Kim, Erik Parner, Amy E Kalkbrenner, Christine Ladd Acosta, M Danielle Fallin, Sherlly Xie, Preben Bo Mortensen, Brian K Lee

**Affiliations:** A.J. Drexel Autism Institute, Drexel University, Philadelphia, PA, USA; The Lundbeck Foundation Initiative for Integrative Psychiatric Research, iPSYCH, Aarhus, Denmark; National Centre for Register-Based Research, Aarhus BSS, Aarhus University, Aarhus, Denmark; Department of Public Health, Aarhus University, Aarhus, Denmark; School of Biomedical Engineering, Science and Health Systems, Drexel University, Philadelphia, PA, USA; University of Wisconsin Milwaukee, Joseph J Zilber College of Public Health, Milwaukee, WI, USA; Department of Epidemiology, Johns Hopkins Bloomberg School of Public Health, Baltimore, MD, USA; Wendy Klag Center for Autism and Developmental Disabilities, Johns Hopkins Bloomberg School of Public Health, Baltimore, MD, USA; Department of Mental Health, Johns Hopkins Bloomberg School of Public Health, Baltimore, MD, USA; Dornsife School of Public Health, Drexel University, Philadelphia, PA, USA; Medtronic, Mounds View, Minnesota, USA; Centre for Integrated Register-based Research, Aarhus University, Aarhus, Denmark; Department of Global Public Health, Karolinska Institutet, Stockholm, Sweden; Department of Epidemiology, Rollins School of Public Health, Emory University, Atlanta, GA USA

**Keywords:** autism, family history, mental disorder, neurologic, cardiometabolic, birth defect, autoimmune, allergy, asthma

## Abstract

The relatively few conditions and family members investigated in autism family health history limits etiologic understanding. For more comprehensive understanding and hypothesis-generation we produced an open-source catalogue of autism associations with family histories of mental, neurologic, cardiometabolic, birth defect, asthma, allergy, and autoimmune conditions. All live births in Denmark, 1980-2012, of Denmark-born parents (1,697,231 births), and their 3-generation family members were followed through April 10, 2017 for each of 90 diagnoses (including autism), emigration or death. Adjusted hazard ratios (aHR) were estimated via Cox regression for each diagnosis-family member type combination, adjusting for birth year, sex, birth weight, gestational age, parental ages at birth, and number of family member types of index person; aHRs also calculated for sex-specific co-occurrence of each disorder. We obtained 6,462 individual family history aHRS across autism overall (26,840 autistic persons; 1.6% of births), by sex, and considering intellectual disability (ID); and 350 individual co-occurrence aHRS. Results are catalogued in interactive heat maps and down- loadable data files: https://ncrr-au.shinyapps.io/asd-riskatlas/ and interactive graphic summaries: https://public.tableau.com/views/ASDPlots_16918786403110/e-Figure5. While primarily for reference material or use in other studies (e.g., meta-analyses), results revealed considerable breadth and variation in magnitude of familial health history associations with autism by type of condition, family member type, sex of the family member, side of the family, sex of the index person, and ID status, indicative of diverse genetic, familial, and non-genetic autism etiologic pathways. Careful attention to sources of autism likelihood in family health history, aided by our open data resource, may accelerate understanding of factors underlying neurodiversity.

**Lay summary:** We calculated the likelihood that a person will be diagnosed with autism if they had a specific family member (e.g, a parent, sibling, grandparent) with a specific mental, neurologic, cardiometabolic, birth defect, asthma, allergy, or autoimmune condition - over 6,000 separate estimates based on 26,840 autistic persons. Results are catalogued in interactive figures and down-loadable data files: https://ncrr-au.shinyapps.io/asd-riskatlas/ and interactive graphic summaries: https://public.tableau.com/views/ASDPlots_16918786403110/e-Figure5. The **st**udy of autism family health history - which varies widely by condition, family member type, sex of the family member, side of the family, sex of the index person, intellectual disability status - may advance understanding of factors underlying neurodiversity.

## Introduction

Strong familial aggregation of autism spectrum disorder (ASD, or autism)^1–7^ was an early clue to autism etiology, suggesting a genetic influence. To date, from comprehensive genome-wide association studies (GWAS) in large samples we can now estimate individual genetic liability for ASD using a polygenic score (PGS),^8^ and detect genetic overlap between ASD and other psychiatric, cognitive and behavioral phenotypes.^8, 9^ For some complex conditions, such as coronary artery disease and schizophrenia, enhanced prediction has been achieved when accounting for both PGS <u>and</u> family history, over PGS alone.^10,11^ Thus, more comprehensive understanding of the role of family health history in autism may complement genetic investigations and accelerate understanding of the origins of neurodiversity.^12^

The investigation of family health history in autism, however, is quite fragmented. While significant autism associations from family histories of mental or non-mental disorders are consistently reported,^13–26^ the typical approach has been to focus on limited numbers of disorders or family members, e.g., parents, siblings – a kind of ‘candidate family history’ approach. Such an approach makes it difficult to assess ASD associations across disorders or family member types which – by achieving a wider perspective - may inform etiologic understanding or additional hypothesis-generation.

Our study goal was to promote efforts towards a more comprehensive understanding of family health history in autism and thereby facilitate future hypothesis-generation by developing an open-source catalogue and associated summary data of autism family health history. Based on a total population cohort design, the catalogue is comprised of: 1) autism associations with 90 mental and non-mental diagnosis categories reported in 3-generation family members distinguished by degree of relatedness, sex, and maternal or paternal side of the family and 2) separate estimates of association per family member type-condition combination: for autism overall, for autism in females and males, and for the autism with intellectual disability (ID) sub-phenotype.

## Methods

We followed STROBE guidelines for study reporting.^27^

### Study population

The study cohort (e-Figure 1) comprised all live births born in Denmark, 1 January 1980 through 31 December 2012, with a valid personal identification number, of known parents who themselves were born in Denmark, and with Medical Birth Register ^28^ information (N= 1,697,231; 82.2% of births). For each study cohort member, we identified twenty, 3-generation family member types (e-Figure 2) via linkage with the Danish Civil Registration Service^29^ (e-Tables 1-2, e-Figure 3).

**Figure 1.**
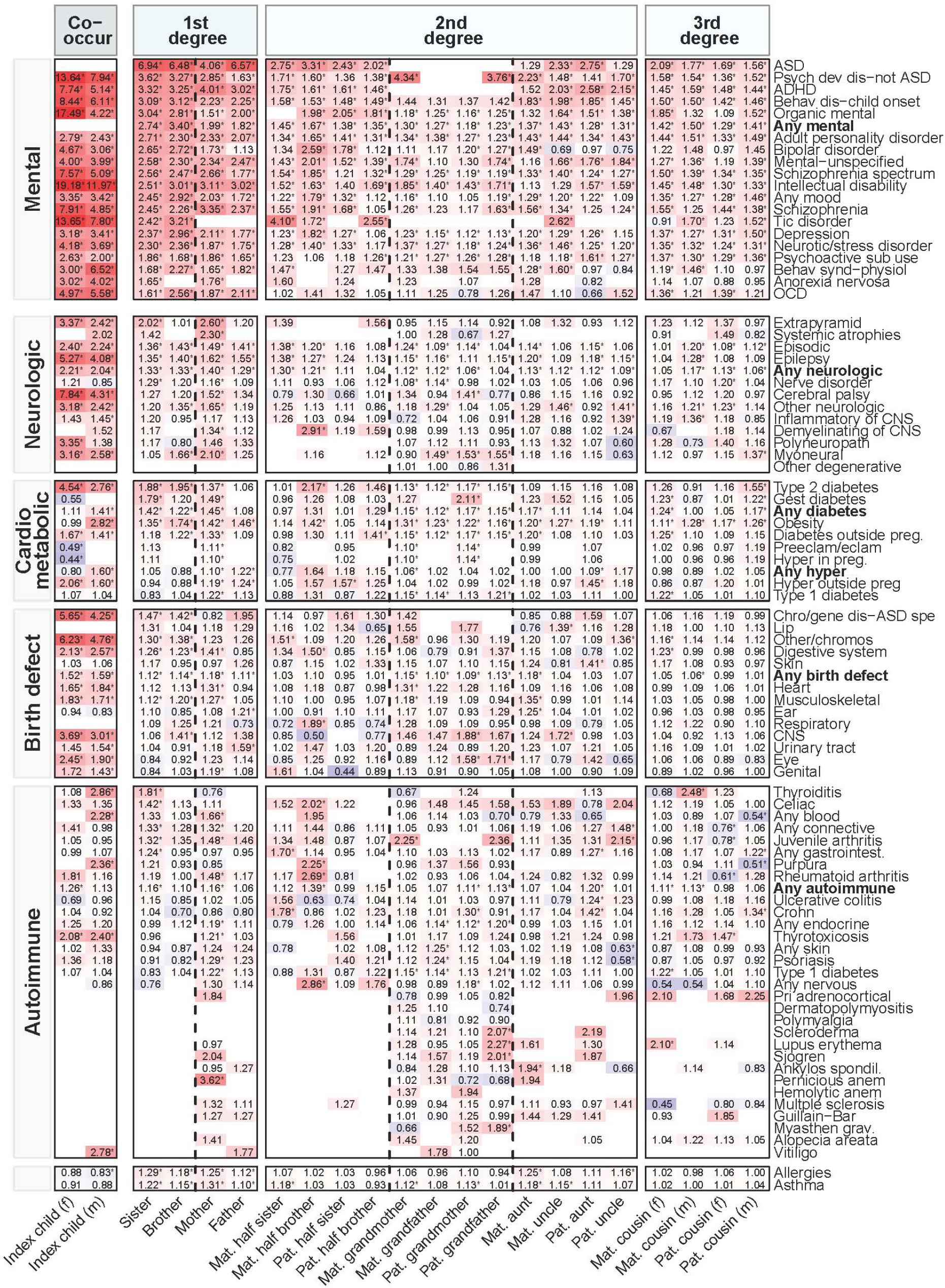
Hazard Ratio^a^ for ASD from family morbidity by diagnosis and family member type. **Hazard Ratio:** the value in a cell of the heat map corresponds to the adjusted Hazard Ratio for ASD estimated from that family member type-morbidity combination. An asterisk (*) next to the cell value indicates that the 95% confidence interval for the adjusted Hazard Ratio excludes the value of 1.0. **Co- occurring condition:** the two co-occurring condition columns correspond to risk of co-occurrence of a specific diagnosis in autistic females and males. **1^st^ degree, 2^nd^ degree, 3^rd^ degree**: each column corresponds to risk for ASD from a specific family member type (such as a full sister) with a specific diagnosis by degree of relatedness. 1^st^ degree: 50% shared genes with index person. 2^nd^ degree: 25% shared genes with index person. 3^rd^ degree: 12.5% shared genes with index person. **Note on blank cells:** a blank cell reflects a sample size of ASD cases of <5 for the specific family member type-diagnosis combination (or <5 ASD cases with the specific co-occurring diagnosis). **Row order:** Within each major diagnosis group, the rows are ordered by diagnosis with the largest aHR (top row) to the smallest aHR (bottom row) from full sisters. **^a^Hazard Ratio (aHR)** adjusted for sex, birth weight, gestational age, parental ages at birth; each aHR estimated using separate baseline ASD --diagnostic rates per birth year. aHRs from full/half siblings, aunts, uncles or cousins also included the number of family members of the relevant type as a covariate.

**Figure 2:**
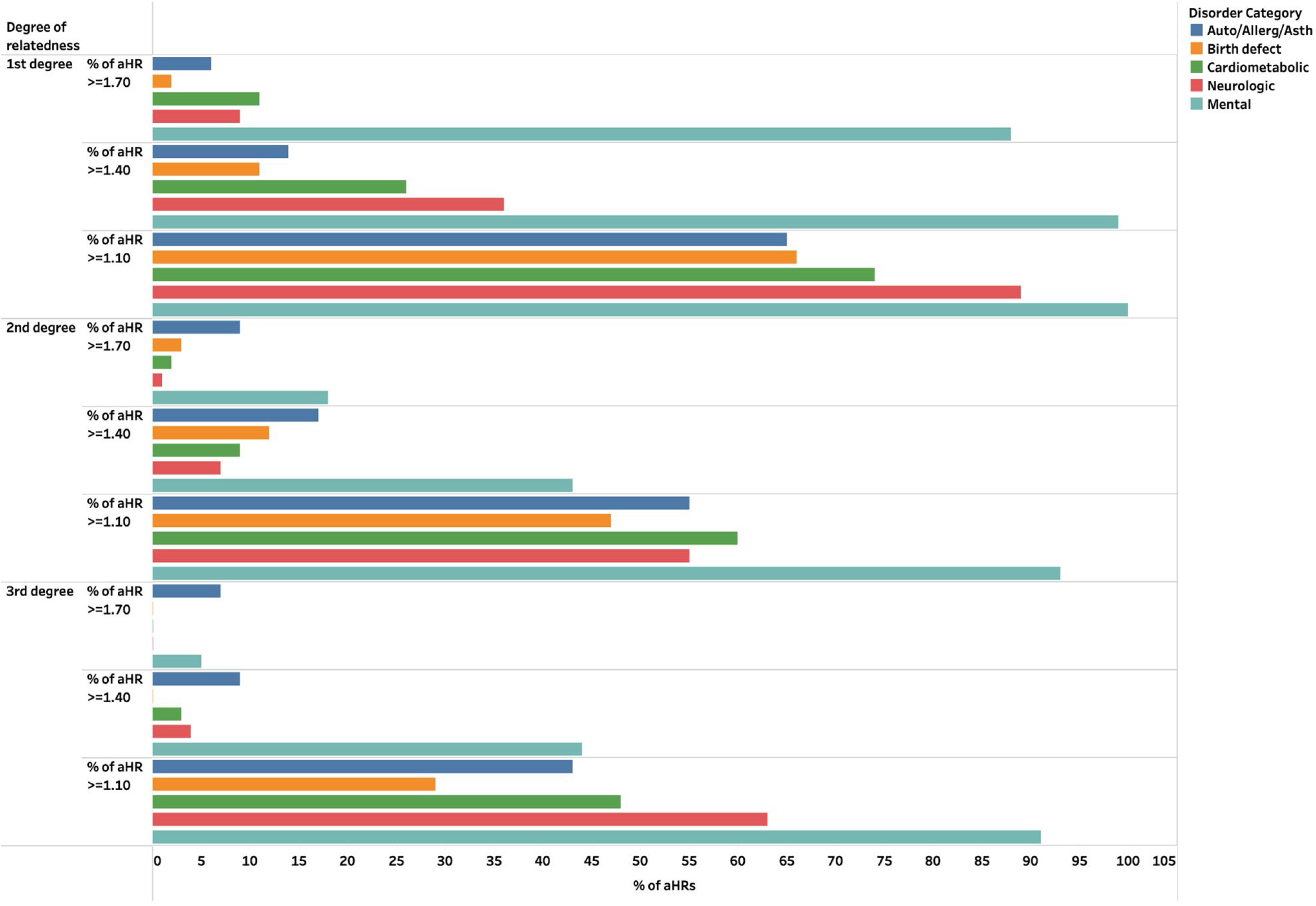
Percent of ASD risk estimates (aHR) by: level of risk, disorder category, in 1st, 2nd or 3rd degree relatives.

**Figure 3:**
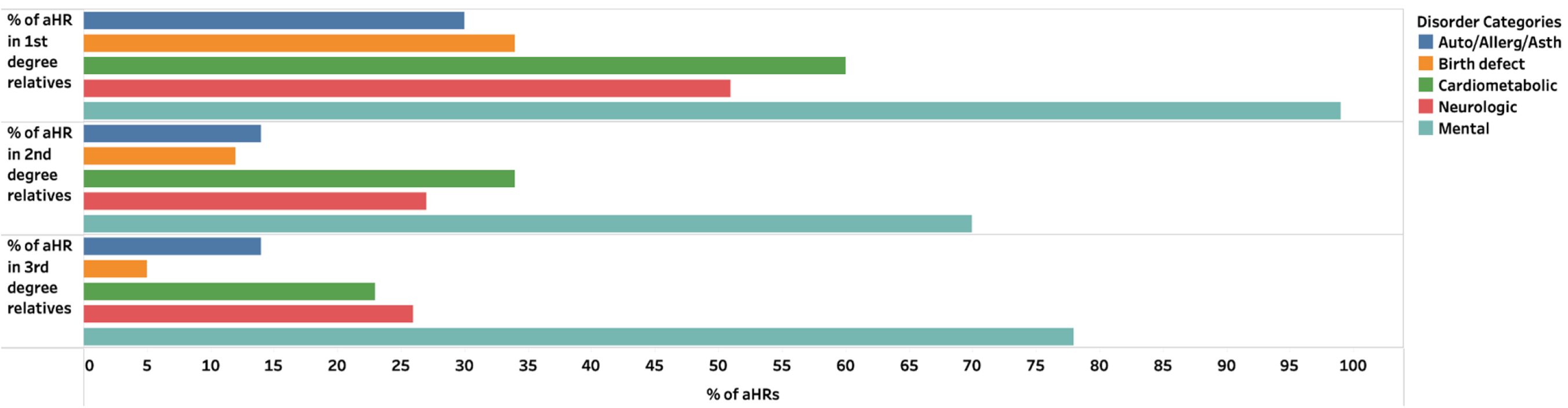
Percent of ASD risk estimates (aHR) with a confidence interval (CI) excludes 1.0 by: disorder category, in 1st, 2nd and 3rd degree relatives.

**Table 1.** Characteristics of persons with and without autism spectrum disorder (ASD) in study cohort.

|  | No ASD |  | ASD |  |  | No ASD |  | ASD |  |  | No ASD |  | ASD |  |
| --- | --- | --- | --- | --- | --- | --- | --- | --- | --- | --- | --- | --- | --- | --- |
|  | n=1,670,391 |  | n=26,840 |  |  | n=1,670,391 |  | n=26,840 |  |  | n=1,670,391 |  | n=26,840 |  |
|  | n | % | n | % |  | n | % | n | % |  | n | % | n | % |
| Male | 851,349 | 51.0 | 19,900 | 74.1 | Birth year |  |  |  |  |  |  |  |  |  |
| Intellectual disability: Yes | 7,697 | 0.5 | 3,342 | 12.5 | 1980 | 51,048 | 3.1 | 153 | 0.6 | 1996 | 54,272 | 3.2 | 1,279 | 4.8 |
| Gestational age (weeks) |  |  |  |  | 1981 | 47,172 | 2.8 | 172 | 0.6 | 1997 | 53,666 | 3.2 | 1,416 | 5.3 |
| ≤36 | 96,874 | 5.9 | 2,040 | 7.7 | 1982 | 46,860 | 2.8 | 236 | 0.9 | 1998 | 52,138 | 3.1 | 1,470 | 5.5 |
| 37-40 | 1,111,458 | 67.6 | 17,467 | 65.6 | 1983 | 45,259 | 2.7 | 248 | 0.9 | 1999 | 517,570 | 3.1 | 1,546 | 5.8 |
| 41+ | 436,989 | 26.6 | 7,112 | 26.7 | 1984 | 45,873 | 2.7 | 292 | 1.1 | 2000 | 51,834 | 3.1 | 1,668 | 6.2 |
| Maternal age (36+ years) | 211,979 | 12.7 | 3,797 | 14.1 | 1985 | 47,435 | 2.8 | 342 | 1.3 | 2001 | 50,275 | 3.0 | 1,496 | 5.6 |
| Paternal age (36+ years) | 430,612 | 25.8 | 7,654 | 28.5 | 1986 | 48,334 | 2.9 | 353 | 1.3 | 2002 | 49,391 | 3.0 | 1,361 | 5.1 |
| Birth weight (grams) |  |  |  |  | 1987 | 48,578 | 2.9 | 399 | 1.5 | 2003 | 50,024 | 3.0 | 1,277 | 4.8 |
| ≤2500 | 88,542 | 5.3 | 1,810 | 6.8 | 1988 | 50,616 | 3.0 | 500 | 1.9 | 2004 | 50,572 | 3.0 | 1,158 | 4.3 |
| 2501-3000 | 210,534 | 12.7 | 3,448 | 12.9 | 1989 | 52,444 | 3.1 | 583 | 2.2 | 2005 | 50,565 | 3.0 | 1,055 | 3.9 |
| 3001-4000 | 1,089,957 | 65.6 | 16,506 | 61.8 | 1990 | 53,925 | 3.2 | 692 | 2.6 | 2006 | 51,207 | 3.1 | 985 | 3.7 |
| 4001+ | 273,317 | 16.4 | 4,929 | 18.5 | 1991 | 53,912 | 3.2 | 788 | 2.9 | 2007 | 50,321 | 3.0 | 857 | 3.2 |
|  |  |  |  |  | 1992 | 56,527 | 3.4 | 877 | 3.3 | 2008 | 50,788 | 3.0 | 656 | 2.4 |
|  |  |  |  |  | 1993 | 55,769 | 3.3 | 970 | 3.6 | 2009 | 48,470 | 2.9 | 561 | 2.1 |
|  |  |  |  |  | 1994 | 57,302 | 3.4 | 1,213 | 4.5 | 2010 | 48,593 | 2.9 | 503 | 1.9 |
|  |  |  |  |  | 1995 | 57,101 | 3.4 | 1,189 | 4.4 | 2011 | 44,783 | 2.7 | 345 | 1.3 |
|  |  |  |  |  |  |  |  |  |  | 2012 | 43,579 | 2.6 | 200 | 0.7 |

### Family health data

The family mental, neurologic, cardiometabolic, birth defect, autoimmune, asthma, and allergy diagnosis categories, and associated individual diagnoses, were selected for study based on previously reported associations with ASD^13–26^ (e-Table 3) and were obtained via linkage with the Psychiatric Central Research and National Patient Registers. ^30, 31^

### Covariates

Child sex, date of birth, birth weight, gestational age, parental ages at birth, death and emigration information was obtained from the Medical Birth Register and Civil Registration Service. By design, the very small proportions of cohort members with missing mother (0.2%), father (0.9%), or birth registry information (0.12%) were excluded (eFigure 1).

### Analytic approach

All persons were followed through April 10, 2017 (cohort members: 5.25 years minimum and 37.25 years maximum follow-up time; family members born before 1980: follow-up began in 1977 with 37.25 years minimum and 40.25 years maximum follow-up time) for medical diagnoses, emigration or death, whichever came first. If more than one sibling, aunt, uncle or cousin had received a specific diagnosis (e.g., two full siblings received an ADHD diagnosis), the earliest diagnosis date was chosen for analysis. For each of the 20 family member types, separate Cox regression models were fitted for each diagnosis. The adjusted hazard ratio (aHR) for ASD was estimated using separate baseline ASD diagnostic rates for each 1-year stratum of birth year, adjusting for sex, birth weight (≤2500g, 2501-3000g, 3001-4000g, 4001+g), gestational age (≤36 weeks, 37-40 weeks, 41+weeks) and parental ages at birth (≤35 years, 36+ years). Models for estimates from full/half siblings, aunts, uncles or cousins also included the number of family members of the relevant type as a covariate. From grandparents, parents, aunts and uncles the diagnosis exposure was based on the family member’s reported diagnosis status (Yes/No per diagnosis) at birth of the index child. For index persons, full/half siblings and cousins the diagnosis exposure was time varying and based on the date of the diagnosis with robust standard errors clustered on the personal identifier of the index child. Separate aHRs were calculated for i) ASD overall, ii) for ASD in females and males and iii) for the subphenotype ASD with ID.

aHRs were also calculated for co-occurrence of each condition in autistic females and males, as well as for autistic persons with ID. aHRs for a co-occurring condition associated with ASD was based on whether ASD had, or had not, been reported for the index person prior to the co-occurring condition.

Sensitivity analyses were carried out to assess the effects of co-occurring ID in family members with a specific health condition on the association between the family health condition and ASD in the index person. These analyses included ID status of the exposing family member as an additional potential confounder variable in models. Similarly, ID status of the autistic person was included as an additional potential confounder variable in separate models to estimate aHRs for disorder co-occurrence in ASD.

No aHR was estimated in any analysis if there were <5 ASD exposed cases per family member type-diagnosis combination (or with a co-occurring condition in the ASD disorder co-occurrence analysis).

This study was approved by the Danish Data Protection Agency and the Danish Health Data Authority. According to Danish law, this secondary use of register data (collected for clinical or administrative purposes), where researchers only have access to pseudo-anonymized information, does not require informed consent.

## Data Availability

The individual-level data that support the findings of this study can only be accessed through Statistics Denmark secure servers where downloading individual level information is prohibited. We obtained approved exports of the necessary summary data needed to generate each aHR and the corresponding 95% confidence interval reported in the study results tables and figures. These data files are accessible here: https://ncrr-au.shinyapps.io/asd-riskatlas/. We prepared a repository (https://github.com/ASDepiteam/ATLAScode) for the data and code used to generate the RShiny webpage for the study which describes the study design and interactive heatmaps of results (adjusted hazard ratios (aHR) calculated via Cox regression models). The data management scripts in the repository incorporate the exported summary data from the study needed to generate each aHR (and corresponding 95% confidence interval) reported in the study results tables, figures and heatmaps. The app.R file is the master file that is used to build and design the interactive features of the RShiny webpage, incorporating the data management scripts to include tables and figures on the webpage.

## Results

Characteristics of autistic (1.6% of the cohort) and non-autistic persons in the study cohort can be found in Table 1.

While we aimed to calculate 9,000 separate aHRs for family history, due to sample size restrictions (no aHR calculation if less than 5 exposed autistic persons per family member-condition combination) we obtained 6,462 aHRs: 1,510 aHRs for autism overall (84% of projected total), 1,436 for autism in males (80% of projected total), 1,136 for autism in females (65% of projected total), 1,424 when adjusting for ID in the family member (79% of projected total) and 956 for ASD with ID (53% of projected total). We also obtained 350 co-occurrence aHRs in autistic males and females (66% of projected total).

To share the large volume of results, our open resource comprises a catalogue of individual estimates in interactive heat maps and downloadable files of the summary data needed to generate each aHR and the corresponding 95% confidence interval (per aHR estimate: logHR, standard error, and n of exposed autistic persons) for autism overall and for associations in males or females, after adjusting for ID in the family member, and for autism with ID (here: https://ncrr-au.shinyapps.io/asd-riskatlas/). Interactive graphic results summaries can be found here: https://public.tableau.com/views/ASDPlots_16918786403110/e-Figure5. While the catalogued results are designed to be used primarily as reference material or for use in other studies (e.g., meta-analyses), the broad scope of results did permit observation of several over-arching patterns briefly highlighted below.

### Family history of mental disorders in autism was more common than non-mental disorders

We observed increased aHRs for autism associated with every <u>mental disorder</u> in family members of all degrees of relatedness in contrast to the more limited frequency of elevated associations among family members with <u>non-mental disorders</u> (Figure 1- heat map; Figure 2-summary plots of % of aHRs at different levels of magnitude by disorder category and degree of family relatedness). Further, 70-99% of the aHRs from <u>mental</u> <u>disorders</u> across all family member types were statistically significant (aHR confidence interval excluded 1.0) while this was less frequently true of the estimates from <u>non-mental disorders</u> (typically 35% or fewer per condition category; Figure 3; e-Figure 4). In autism overall, all aHRs were < 3.0 (Figure 1), except for a 1^st^ degree family history of ASD (aHRs 4-7 for ASD associated with ASD diagnosed in mother, father, full brother, full sister) or ADHD, ID, other developmental disorders, organic mental disorders, tic disorders, schizophrenia, or pernicious anemia, as well as with maternal grandparents diagnosed with other developmental disorders, or a maternal half-sister diagnosed with tic disorders (aHRs 3-4).

### Differences in ASD associations by sex of the family member or side of the family were common in non-mental disorders

Among <u>non-mental disorders</u> the proportions of aHRs at different magnitudes, or proportions of aHRs that were statistically significant, were higher with an affected mother than father (e-Figures 5-6). These proportions varied also among full siblings by sex (e.g., differences between brothers and sisters with autoimmune disorders in magnitude of aHRs or proportions of aHRS that were statistically significant; e-Figures 7-8) and among half siblings by side of the family (e.g., higher proportions from affected half siblings on the maternal than paternal side of the family with neurologic, birth defect or autoimmune disorders; e-Figures 9-10). aHRs from cardiometabolic conditions in grandparents of both sexes and sides of the family were primarily in the range of aHRs 1.10-1.39 (e-Figures 11 and 13) and over 70% of these estimates were also statistically significant (e-Figures 12 and 14). For birth defects, the proportions of elevated ASD risks or of estimates that were statistically significant were somewhat higher on the maternal than paternal side of the family and in female more than male family members (e-Figures 15-18). For asthma, all ASD aHRs fell below aHR 1.40 but the proportions of all estimates that fell in the range of aHR 1.10-1.39 were higher with affected family members on the maternal side of the family (50% versus 30% on paternal side) or were female (60% versus 30% among males) as were the proportions of estimates that were statistically significant (50% on maternal side versus 20% on paternal side; 60% in female family members versus 30% in males).

### ASD family history patterns in males and females separately were similar to ASD overall (e-Figures 19-20)

As with ASD overall, there was a preponderance of elevated ASD aHRs from <u>mental</u> versus <u>non-mental disorders</u> (e-Figure 21) and about 60%-70% of <u>mental disorder</u> estimates were statistically significant compared to about 30% or fewer of <u>non-mental disorder</u> estimates (e-Figure 22). With <u>non-mental disorders</u> the proportions of elevated ASD aHRs in both males and females, or proportions of estimates that were statistically significant, were higher with an affected mother than father (e-Figures 23-26).

### Presence of ID impacts select aHRs

When accounting for co-occurring ID in the family member, the magnitude of the aHR for ASD per diagnosis in the family member generally either remained the same or was attenuated compared to results when not accounting for ID (e-Figures 27-46). For the ASD with ID outcome, unlike in ASD overall, there was no preponderance of elevated associations with maternal neurologic, birth defect or autoimmune disorders (e-Figures 47-48) compared to paternal associations (e-Figures 5-6).

### Co-occurrence versus family history patterns

As with family history, there were elevated aHRs for co-occurrence of every <u>mental disorder</u> in autistic males and females while elevated aHRs of co-occurrence of <u>non-mental disorders</u> were less frequent (e-Figures 49-50); when there were significant co-occurrence aHRs in both sexes, the aHR in autistic females was in most instances higher than in males (e-Figure 51). If a condition was associated with both a significantly elevated ASD aHR for co-occurrence <u>and</u> elevated familial aHRs, the co-occurrence aHR was generally higher than the familial aHR (e-Figures 52-53). The aHR of co-occurrence of a second specific diagnosis in autistic persons typically was attenuated when accounting for co-occurring ID in the autistic person, especially in autistic females, compared to results not accounting for ID (e-Figures 54-55). Compared to autism overall, in autistic persons with ID, for some disorders the co-occurrence aHR was higher (e.g., epilepsy, CNS birth defects, Type 2 diabetes) while in others the aHR was lower (e.g., most mental disorders) (e-Figures 57-58). As seen in ASD overall, there were higher co-occurrence aHRs for schizophrenia, epilepsy, Type 2 diabetes, and chromosomal defects in autistic females with ID than autistic males with ID (e- Figure 59).

## Discussion

Our comprehensive approach revealed the considerable breadth and depth of the range of disorders in families associated with ASD occurrence and all results are fully available for further examination or use in other studies via open access. Many individual findings are consistent with the literature, yet the ability to also have an over- arching view yielded perspectives into potential sources and patterns of family medical history associated with ASD to be explored in further studies designed for specific hypothesis-testing.

For example, we observed greater frequency of significant aHRs for ASD in the index person associated with neurologic, cardiometabolic, birth defect or autoimmune disorders in mothers compared to fathers. We also observed greater frequency of elevated aHRs arising from maternal than paternal half-siblings with neurologic, birth defect or autoimmune disorders – maternal half siblings (sharing the same mother but not father) are more likely to share prenatal environmental factors, and possibly postnatal environments, than paternal half siblings. ASD, notably, may be only one of many developmental effects of a specific maternal condition. For example, besides increased ASD risk, maternal prenatal diabetes or obesity increases offspring risk for birth defects^32–34^ and maternal prenatal autoimmune disease increases risk for adverse pregnancy and developmental outcomes.^35–38^ Thus, the elevated aHRs for ASD in our data associated with a wide array of non-mental conditions in a mother, full sib or maternal half sib may reflect a wide profile of different offspring outcomes linked via diverse pathways to specific maternal prenatal conditions. This hypothesis could be tested in future family-based designs by examining diverse outcome patterns within sibships associated with specific maternal conditions.

Second, significant ASD aHRs associated with grandparents (of both sexes and sides of the family) with cardiometabolic conditions to our knowledge has not been previously reported and may warrant further investigation of the potential for shared genetic liability between these disorders and ASD or possibly adverse lifestyles shared across generations that influence non-genetic prenatal risks. ^39, 40^

Third, a greater likelihood of autoimmune disorders ^41, 42^ and adult asthma ^43–45^ to occur in females than males that has been reported in other studies may partly explain the relatively greater frequency of elevated ASD aHRs in our data arising from autoimmune disorders in mothers and sisters (versus fathers and brothers) or from asthma in female family members (mostly on the maternal side of the family) (i.e., these conditions, as ‘exposures’, are more common in females than males and thereby we may more likely detect an association in female than male family members). With respect to asthma, however, the adult asthma preponderance in females seen in other studies may be limited to non-allergic asthma. ^46^ Thus, further study of whether the asthma in female family members and ASD associations observed here are centered on non-allergic asthma may render clues to potential shared pathogenic mechanisms (e.g., sex hormone or obesity effects) ^46^ linking asthma and ASD in the same family. In contrast to asthma or autoimmune disorders, birth defects are more common among males,^47^ yet we observed more frequent elevated ASD aHRs associated with birth defects in female family members or on the maternal side of the family – a sex-related difference which may warrant further scrutiny also.

We observed that the aHR for ASD from a disorder in a family member after accounting for co-occurring ID in the family member most often involved attenuation of the aHR for ASD in the index person which is consistent with the hypothesis that genetic risk for autism may not be specific for autism alone, but also overlap other conditions such as ID ^48^; thus the apparent elevated ASD aHR associated with the disorder in the affected family member may have been confounded by the co-occurrence of ID in the family member. Another ID effect on ASD was seen in the aHRS for the ASD with ID outcome: unlike in ASD overall, there were no differences observed in the aHRs for ASD with ID associated with mothers versus fathers with neurologic, birth defect or autoimmune conditions. The latter pattern suggests that there may be prenatal influences of maternal neurologic, birth defect or autoimmune conditions on ASD overall but not on ASD with ID, specifically.

Finally, while our observed co-occurrence likelihoods in persons with ASD are consistent with previous studies^25, 26, 49–52^, we further observed risk differences by sex (e.g., higher female co-occurrence risk across most conditions) and the impact of ID on ASD co-occurrence risk, especially in autistic females.

The strength of our approach was the systematic, population-based estimation of ASD associations from a single population and methodology across multiple disorder and family member type combinations. This approach, plus provision of an open, interactive display of estimates for each disorder-ASD association, provides important context for comparison, interpretation, and further hypothesis generation for future studies. We chose to not correct for multiple testing since our aims were to produce a data resource for hypothesis-generation and baseline comparison with other studies. Additionally, more complex modeling or confounder adjustment per condition was beyond the scope of the analysis and each result will need closer scrutiny. Some associations based on small sample sizes may be spurious. Since general practitioners do not report to Danish registries, our data are relevant to conditions diagnosed by specialists. Lastly, we focused on pathways not acting via factors impacting birth weight and gestational age.

## Conclusion

Our investigation of family medical history in ASD revealed considerable breadth and variation in patterns of association suggestive of diverse genetic, familial and non-genetic ASD etiologic pathways. Family medical history complements genetic measures,^12^ perhaps by encompassing unidentified genetic (e.g., not captured by current PGS) and non-genetic components.^53^ Thus, more careful attention to mechanisms of autism likelihood encompassed in family medical history, in addition to genetics,^54, 55^ may accelerate understanding of factors underlying neurodiversity. We hope our findings in an open data resource will stimulate such hypothesis testing.

## Funding Acknowledgements

The study was supported by the Lundbeck Foundation (iPSYCH, Grant numbers R102-A9118 and R155-2014- 1724), the National Institute of Environmental Health Sciences of the National Institutes of Health under Award No. R01ES026993, and the National Institute of Neurologic Disorders and Stroke of the National Institutes of Health under Award No. R01 NS131433. The funding sponsors had no role in the study design; in the collection, analysis, and interpretation of data; in the writing of the report; and in the decision to submit the paper for publication. The content is solely the responsibility of the authors and does not necessarily represent the official views of the National Institutes of Health.

## Competing Interest Statement

The authors have no competing interests to declare.

## Notes

### Competing Interest Statement

The authors have declared no competing interest.

### Summary of Updates

Abstract and main text shortened to accommodate target journal limits; study objective text modified to emphasize study product as a catalogue of findings and open data resource; 2 figures removed

