## Supplemental tables and figures for "3-generation family histories of mental, neurologic, cardiometabolic, birth defect, asthma, allergy, and autoimmune conditions associated with autism: an open-source catalogue of findings"

<u>List of supplementary tables and figures</u>

**eTable 1. Number and percent of study cohort members with a linkage to at least one person of each specific family member type**

**eTable 2. Mean number of cousins, aunts, uncles and siblings per person in the study cohort**

**eTable 3. Diagnosis identification**

**eFigure 1. Selection of study cohort**

**eFigure 2. Overview of family member types and diagnosis categories for analysis and risks to be estimated**

**eFigure 3. Percent of study cohort members with linkages to siblings, grandparents, aunts, uncles and cousins by birth year**

**eFigure 4. Risk for ASD (aHR) from family morbidity by diagnosis and family member type, statistically significant estimates only (aHR confidence interval excludes 1.0)**

**To see eFigures 5 through 18, eFigures 21 through 55, or eFigures 57 through 59, click on accompanying hyperlink with a specific eFigure. To reach website with all eFigures, click on this link** https://public.tableau.com/views/ASDPlots_16918786403110/e-Figure5 **and select appropriate tab on webpage (e.g., tab for eFigure 5).**

**eFigure 5: Risk for ASD (aHR) from diagnosis of a condition in mothers or fathers by: level of risk, disorder category**

**https://public.tableau.com/shared/YFCHW64TN?:display_count=n&:origin=viz_share_link**

**eFigure 6: Percent of ASD risk estimates (aHR) from diagnosis of a condition in mothers or fathers with a confidence interval (CI) excluding 1.0 by: disorder category**

**https://public.tableau.com/views/ASDPlots_16918786403110/e-Figure6?:language=en-US&:display_count=n&:origin=viz_share_link**

**eFigure 7: Risk for ASD (aHR) from diagnosis of a condition in full sisters or full brothers by: level of risk, disorder category**

**https://public.tableau.com/views/ASDPlots_16918786403110/e-Figure7?:language=en-US&:display_count=n&:origin=viz_share_link**

**eFigure 8: Percent of ASD risk estimates (aHR) from diagnosis of a condition in full sisters or full brothers with a confidence interval (CI) excluding 1.0 by: disorder category**

https://public.tableau.com/views/ASDPlots_16918786403110/e-Figure8?:language=en-US&:display_count=n&:origin=viz_share_link

**e-Figure 9: Risk for ASD (aHR) from diagnosis of a condition in paternal or maternal half siblings by: level of risk, disorder category**

https://public.tableau.com/views/ASDPlots_16918786403110/e-Figure9?:language=en-US&:display_count=n&:origin=viz_share_link

**e-Figure 10: Percent of ASD risk estimates (aHR) from diagnosis of a condition in paternal or maternal half siblings with a confidence interval (CI) excluding 1.0 by: disorder category**

https://public.tableau.com/views/ASDPlots_16918786403110/e-Figure10?:language=en-US&:display_count=n&:origin=viz_share_link

**e-Figure 11: Risk for ASD (aHR) from diagnosis of a condition in paternal or maternal grandparents by: level of risk, disorder category**
https://public.tableau.com/views/ASDPlots_16918786403110/e-Figure11?:language=en-US&:display_count=n&:origin=viz_share_link

**e-Figure 12: Percent of ASD risk estimates (aHR) from diagnosis of a condition in paternal or maternal grandparents with a confidence interval (CI) excluding 1.0 by: disorder category**

https://public.tableau.com/views/ASDPlots_16918786403110/e-Figure12?:language=en-US&:display_count=n&:origin=viz_share_link

**e-Figure 13: Risk for ASD (aHR) from diagnosis of a condition in grandfathers or grandmothers by: level of risk, disorder category**

https://public.tableau.com/views/ASDPlots_16918786403110/e-Figure13?:language=en-US&:display_count=n&:origin=viz_share_link

**e-Figure 14: Percent of ASD risk estimates (aHR) from diagnosis of a condition in grandfathers or grandmothers with a confidence interval (CI) excluding 1.0 by: disorder category**

https://public.tableau.com/views/ASDPlots_16918786403110/e-Figure14?:language=en-US&:display_count=n&:origin=viz_share_link

**e-Figure 15: Risk for ASD (aHR) from diagnosis of a condition in the paternal or maternal side of the family by: level of risk, disorder category**

**https://public.tableau.com/views/ASDPlots_16918786403110/e-Figure15?:language=en-US&:display_count=n&:origin=viz_share_link**

**e-Figure 16: Percent of ASD risk estimates (aHR) from diagnosis of a condition in the paternal or maternal side of the family with a confidence interval (CI) excluding 1.0 by: disorder category**

**https://public.tableau.com/views/ASDPlots_16918786403110/e-Figure16?:language=en-US&:display_count=n&:origin=viz_share_link**

**e-Figure 17: Risk for ASD (aHR) from diagnosis of a condition in male or female family members by: level of risk, disorder category**

**https://public.tableau.com/views/ASDPlots_16918786403110/e-Figure17?:language=en-US&:display_count=n&:origin=viz_share_link**

**e-Figure 18: Percent of ASD risk estimates (aHR) from diagnosis of a condition in male or female family members with a confidence interval (CI) excluding 1.0 by: disorder category**

**https://public.tableau.com/views/ASDPlots_16918786403110/e-Figure18?:language=en-US&:display_count=n&:origin=viz_share_link**

**eFigure 19. Risk for ASD (aHR) in males from family morbidity by diagnosis and family member type (see efigure below)**

**eFigure 20. Risk for ASD (aHR) in females from family morbidity by diagnosis and family member type (see efigure below)**

**e-Figure 21: Risk for ASD (aHR) in males or females from diagnosis of a condition across 1st, 2nd and 3rd degree family members by: level of risk, disorder category**

**https://public.tableau.com/views/ASDPlots_16918786403110/e-Figure21?:language=en-US&:display_count=n&:origin=viz_share_link**

**e-Figure 22: Percent of ASD risk estimates (aHR) in males or females from diagnosis of a condition across 1st, 2nd, and 3rd degree family members with a confidence interval (CI) excluding 1.0 by: disorder category**

**https://public.tableau.com/views/ASDPlots_16918786403110/e-Figure22?:language=en-US&:display_count=n&:origin=viz_share_link**

**e-Figure 23: Risk for ASD (aHR) in males from diagnosis of a condition in mothers or fathers by: level of risk, disorder category**

**https://public.tableau.com/views/ASDPlots_16918786403110/e-Figure23?:language=en-US&:display_count=n&:origin=viz_share_link**

**e-Figure 24: Percent of ASD risk estimates (aHR) in males from diagnosis of a condition in mothers or fathers with a confidence interval (CI) excluding 1.0 by: disorder category**

https://public.tableau.com/views/ASDPlots_16918786403110/e-Figure24?:language=en-US&:display_count=n&:origin=viz_share_link

**e-Figure 25: Risk for ASD (aHR) in females from diagnosis of a condition in mothers or fathers by: level of risk, disorder category**

https://public.tableau.com/views/ASDPlots_16918786403110/e-Figure25?:language=en-US&:display_count=n&:origin=viz_share_link

**e-Figure 26: Percent of ASD risk estimates (aHR) in females from diagnosis of a condition in mothers or fathers with a confidence interval (CI) excluding 1.0 by: disorder category**

https://public.tableau.com/views/ASDPlots_16918786403110/e-Figure26?:language=en-US&:display_count=n&:origin=viz_share_link

**e-Figure 27: Risk for ASD (aHR) from a maternal diagnosis overall versus after adjusting for Intellectual Disability (ID) in the mother by: specific diagnosis**

https://public.tableau.com/views/ASDPlots_16918786403110/e-Figure27?:language=en-US&:display_count=n&:origin=viz_share_link

**e-Figure 28: Risk for ASD (aHR) from a paternal diagnosis overall versus after adjusting for Intellectual Disability (ID) in the father by: specific diagnosis**

https://public.tableau.com/views/ASDPlots_16918786403110/e-Figure28?:language=en-US&:display_count=n&:origin=viz_share_link

**e-Figure 29: Risk for ASD (aHR) from a diagnosis in the sister overall versus after adjusting for Intellectual Disability (ID) in the sister by: specific diagnosis**

https://public.tableau.com/views/ASDPlots_16918786403110/e-Figure29?:language=en-US&:display_count=n&:origin=viz_share_link

**e-Figure 30: Risk for ASD (aHR) from a diagnosis in the brother overall versus after adjusting for Intellectual Disability (ID) in the brother by: specific diagnosis**

https://public.tableau.com/views/ASDPlots_16918786403110/e-Figure30?:language=en-US&:display_count=n&:origin=viz_share_link

**e-Figure 31: Risk for ASD (aHR) from a maternal half sister diagnosis overall versus after adjusting for Intellectual Disability (ID) in the maternal half sister by: specific diagnosis**

https://public.tableau.com/views/ASDPlots_16918786403110/e-Figure31?:language=en-US&:display_count=n&:origin=viz_share_link

**e-Figure 32: Risk for ASD (aHR) from a maternal half brother diagnosis overall versus after adjusting for Intellectual Disability (ID) in the maternal half brother by: specific diagnosis**

https://public.tableau.com/views/ASDPlots_16918786403110/e-Figure32?:language=en-US&:display_count=n&:origin=viz_share_link

**e-Figure 33: Risk for ASD (aHR) from a paternal half sister diagnosis overall versus after adjusting for Intellectual Disability (ID) in the paternal half sister by: specific diagnosis**

https://public.tableau.com/views/ASDPlots_16918786403110/e-Figure33?:language=en-US&:display_count=n&:origin=viz_share_link

**e-Figure 34: Risk for ASD (aHR) from a paternal half brother diagnosis overall versus after adjusting for Intellectual Disability (ID) in the paternal half brother by: specific diagnosis**

https://public.tableau.com/views/ASDPlots_16918786403110/e-Figure34?:language=en-US&:display_count=n&:origin=viz_share_link

**e-Figure 35: Risk for ASD (aHR) from a maternal grandmother diagnosis overall versus after adjusting for Intellectual Disability (ID) in the maternal grandmother by:**
https://public.tableau.com/views/ASDPlots_16918786403110/e-Figure35?:language=en-US&:display_count=n&:origin=viz_share_link

**e-Figure 36: Risk for ASD (aHR) from a maternal grandfather overall versus after adjusting for Intellectual Disability (ID) in the maternal grandfather by: specific diagnosis**

https://public.tableau.com/views/ASDPlots_16918786403110/e-Figure36?:language=en-US&:display_count=n&:origin=viz_share_link

**e-Figure 37: Risk for ASD (aHR) from a paternal grandmother diagnosis overall versus after adjusting for Intellectual Disability (ID) in the paternal grandmother by: specific diagnosis**

https://public.tableau.com/views/ASDPlots_16918786403110/e-Figure37?:language=en-US&:display_count=n&:origin=viz_share_link

**e-Figure 38: Risk for ASD (aHR) from a paternal grandfather diagnosis overall versus after adjusting for Intellectual Disability (ID) in the paternal grandfather by: specific diagnosis**

https://public.tableau.com/views/ASDPlots_16918786403110/e-Figure38?:language=en-US&:display_count=n&:origin=viz_share_link

**e-Figure 39: Risk for ASD (aHR) from a maternal aunt diagnosis overall versus after adjusting for Intellectual Disability (ID) in the maternal aunt by: specific diagnosis**

https://public.tableau.com/views/ASDPlots_16918786403110/e-Figure39?:language=en-US&:display_count=n&:origin=viz_share_link

**e-Figure 40: Risk for ASD (aHR) from a maternal uncle diagnosis overall versus after adjusting for Intellectual Disability (ID) in the maternal uncle by: specific diagnosis**

https://public.tableau.com/views/ASDPlots_16918786403110/e-Figure40?:language=en-US&:display_count=n&:origin=viz_share_link

**e-Figure 41: Risk for ASD (aHR) from a paternal aunt diagnosis overall versus after adjusting for Intellectual Disability (ID) in the paternal aunt by: specific diagnosis**

https://public.tableau.com/views/ASDPlots_16918786403110/e-Figue41?:language=en-US&:display_count=n&:origin=viz_share_link

**e-Figure 42: Risk for ASD (aHR) from a paternal uncle diagnosis overall versus after adjusting for Intellectual Disability (ID) in the paternal unclde by: specific diagnosis**

https://public.tableau.com/views/ASDPlots_16918786403110/e-Figure42?:language=en-US&:display_count=n&:origin=viz_share_link

**e-Figure 43: Risk for ASD (aHR) from a maternal female cousin diagnosis overall versus after adjusting for Intellectual Disability (ID) in the maternal female cousin by: specific diagnosis**

https://public.tableau.com/views/ASDPlots_16918786403110/e-Figure43?:language=en-US&:display_count=n&:origin=viz_share_link

**e-Figure 44: Risk for ASD (aHR) from a maternal male cousin diagnosis overall versus after adjusting for Intellectual Disability (ID) in the maternal male cousin by: specific diagnosis**

https://public.tableau.com/views/ASDPlots_16918786403110/e-Figure44?:language=en-US&:display_count=n&:origin=viz_share_link

**e-Figure 45: Risk for ASD (aHR) from a paternal female cousin diagnosis overall versus after adjusting for Intellectual Disability (ID) in the paternal female cousin by: specific diagnosis**

https://public.tableau.com/views/ASDPlots_16918786403110/e-Figure45?:language=en-US&:display_count=n&:origin=viz_share_link

**e-Figure 46: Risk for ASD (aHR) from a paternal male cousin diagnosis overall versus after adjusting for Intellectual Disability (ID) in the paternal male cousin by: specific diagnosis**

https://public.tableau.com/views/ASDPlots_16918786403110/e-Figure46?:language=en-US&:display_count=n&:origin=viz_share_link

**e-Figure 47: Risk for ASD with Intellectual Disability (aHR) from diagnosis of a condition in mothers or fathers by: level of risk, disorder category**

https://public.tableau.com/views/ASDPlots_16918786403110/e-Figure47?:language=en-US&:display_count=n&:origin=viz_share_link

**e-Figure 48: Percent of risk estimates (aHR) for ASD with Intellectual Disability from diagnosis of a condition in mothers or fathers with a confidence interval (CI) excluding 1.0 by: disorder category**

https://public.tableau.com/views/ASDPlots_16918786403110/e-Figure48?:language=en-US&:display_count=n&:origin=viz_share_link

**e-Figure 49: Risk of co-occurrence of a second condition in autistic males and females by: level of risk, disorder category**

https://public.tableau.com/views/ASDPlots_16918786403110/e-Figure49?:language=en-US&:display_count=n&:origin=viz_share_link

**e-Figure 50: Percent of risk estimates (aHR) for co-occurrence of a second condition in autistic males or females with a confidence interval (CI) excluding 1.0 by: disorder category**

https://public.tableau.com/views/ASDPlots_16918786403110/e-Figure50?:language=en-US&:display_count=n&:origin=viz_share_link

**e-Figure 51: Risk (aHR) of co-occurrence of a second condition in autistic males versus autistic females by: specific condition**

https://public.tableau.com/views/ASDPlots_16918786403110/e-Figure51?:language=en-US&:display_count=n&:origin=viz_share_link

**e-Figure 52: Risk (aHR) of co-occurrence of a second condition in autistic females versus ASD risk in females from diagnosis of same condition in a 1st degree family member* by: specific condition**

https://public.tableau.com/views/ASDPlots_16918786403110/e-Figure52?:language=en-US&:display_count=n&:origin=viz_share_link

**e-Figure 53: Risk (aHR) of co-occurrence of a second condition in autistic males versus ASD risk in males from diagnosis of same condition in a 1st degree family member* by: specific condition**

https://public.tableau.com/views/ASDPlots_16918786403110/e-Figure53?:language=en-US&:display_count=n&:origin=viz_share_link

**e-Figure 54: Risk (aHR) of co-occurrence of a second condition in autistic males overall versus risk of co-occurrence after adjusting for Intellectual Disability in autistic male by: specific condition**

https://public.tableau.com/views/ASDPlots_16918786403110/e-Figure54?:language=en-US&:display_count=n&:origin=viz_share_link

**e-Figure 55: Risk (aHR) of co-occurrence of a second condition in autistic females overall versus risk of co-occurrence after adjusting for Intellectual Disability in autistic female by: specific condition**
https://public.tableau.com/views/ASDPlots_16918786403110/e-Figure55?:language=en-US&:display_count=n&:origin=viz_share_link

**eFigure 56. Risk for ASD (aHR) with intellectual disability (ID) from family morbidity by diagnosis and family member type (see efigure below)**

**e-Figure 57: Risk (aHR) of co-occurrence of a second condition in autistic males overall versus risk of co-occurrence in autistic males with Intellectual Disability by: specific condition**

https://public.tableau.com/views/ASDPlots_16918786403110/e-Figure57?:language=en-US&:display_count=n&:origin=viz_share_link

**e-Figure 58: Risk (aHR) of co-occurrence of a second condition in autistic females overall versus risk of co-occurrence in autistic females with Intellectual Disability by: specific condition**

https://public.tableau.com/views/ASDPlots_16918786403110/e-Figure58?:language=en-US&:display_count=n&:origin=viz_share_link

**e-Figure 59: Risk (aHR) of co-occurrence of a second condition in autistic males with Intellectual Disability versus risk of co-occurrence in autistic females with Intellectual Disability by: specific condition**

https://public.tableau.com/views/ASDPlots_16918786403110/e-Figure59?:language=en-US&:display_count=n&:origin=viz_share_link

**eTable 1. Number and percent of study cohort members with a linkage to at least one person of each specific family member type**

| | No ASD *n=1,670,601* | | ASD *n=26,843* | |
|---|---|---|---|---|
| **Family member type** | **n** | **%** | **n** | **%** |
| **Maternal** | | | | |
| grandmother | 1,524,592 | 91.3 | 25,816 | 96.2 |
| grandfather | 1,502,402 | 89.9 | 25,472 | 94.9 |
| uncle | 855,646 | 51.2 | 14,004 | 52.2 |
| aunt | 785,681 | 47.0 | 13,282 | 49.5 |
| Female cousin | 896,791 | 53.7 | 14,537 | 54.2 |
| Male cousin | 912,658 | 54.6 | 14,915 | 55.6 |
| Half sister | 155,640 | 9.3 | 3,415 | 12.7 |
| Half brother | 163,582 | 9.8 | 3,385 | 12.6 |
| **Paternal** | | | | |
| grandmother | 1,473,254 | 88.2 | 25,057 | 93.4 |
| grandfather | 1,447,639 | 86.7 | 24,644 | 91.8 |
| uncle | 812,483 | 48.6 | 13,522 | 50.4 |
| aunt | 769,523 | 46.1 | 12,746 | 47.5 |
| Female cousin | 877,953 | 52.6 | 14,322 | 53.4 |
| Male cousin | 893,461 | 53.5 | 14,689 | 54.7 |
| Half sister | 181,295 | 10.9 | 3,588 | 13.4 |
| Half brother | 187,773 | 11.2 | 3,736 | 13.9 |
| **Full sibling** | 1,397,370 | 83.7 | 20,638 | 76.9 |
| **Full brother** | 865,136 | 51.8 | 12,588 | 46.9 |
| **Full sister** | 827,446 | 49.5 | 12,169 | 45.3 |

**eTable 2. Mean number of cousins, aunts, uncles and siblings per person in the study cohort**

| | No ASD *n=1,670,391* Number per person | | ASD *n=26,840* Number per person | |
|---|---|---|---|---|
| Family member type | Mean | standard deviation | Mean | standard deviation |
| **Maternal** | | | | |
| female cousins | 1.13 | 1.52 | 1.12 | 1.45 |
| male cousins | 1.19 | 1.57 | 1.18 | 1.50 |
| aunts | 0.63 | 0.81 | 0.66 | 0.80 |
| uncles | 0.70 | 0.83 | 0.71 | 0.84 |
| half sisters | 0.12 | 0.39 | 0.16 | 0.45 |
| half brothers | 0.12 | 0.41 | 0.16 | 0.46 |
| **Paternal** | | | | |
| female cousins | 1.13 | 1.52 | 1.13 | 1.55 |
| male cousins | 1.19 | 1.59 | 1.19 | 1.58 |
| aunts | 0.61 | 0.78 | 0.63 | 0.79 |
| uncles | 0.68 | 0.86 | 0.71 | 0.86 |
| half sisters | 0.14 | 0.44 | 0.17 | 0.50 |
| half brothers | 0.15 | 0.46 | 0.18 | 0.51 |
| Full siblings | 1.25 | 0.88 | 1.13 | 0.89 |
| Full sisters | 0.61 | 0.70 | 0.54 | 0.68 |
| Full brothers | 0.65 | 0.73 | 0.58 | 0.72 |

**eTable 3. Diagnosis identification**

**Information on mental or behavioral disorder diagnoses was obtained from the Psychiatric Central Research Register. Diagnoses of neurologic conditions, cardiometabolic conditions, birth defects, autoimmune conditions, allergies or asthma were obtained from the Danish National Patient Register. Both registers hold all inpatient diagnoses given at discharge from somatic and psychiatric wards in all hospitals since 1977 (since 1969 for the Psychiatric Register); since 1995 both also hold all discharge diagnoses from outpatient and emergency room contacts. The International Classification of Diseases - Eighth Revision (ICD-8) was used for diagnostic reporting to the registers through 1993 and thereafter using ICD-10. Persons suspected of ASD or other mental or behavioral disorders are referred (e.g., by general practitioners or school psychologists) to a psychiatric department for a multidisciplinary evaluation and are assigned a diagnosis by a psychiatrist; Danish health care is universal and free of charge. All diagnoses are reported to the Psychiatric Register once a diagnosis is established and without regard to the need for treatment or educational provisions. Registry reporting is done only by psychiatrists following mandatory training in the use of the ICD.**
***Summary diagnosis categories are noted in italics***

| | ICD 8 codes | ICD 10 codes |
|---|---|---|
| Mental disorders | | |
| ASD | 299.00-299.03 | F84.0, F84.1, F84.5, F84.8, F84.9 |
| *Any mental* | 290-315 | F00-F99 |
| organic mental | 290, 292, 293, 294, 309 | F00-F09 |
| psychoactive sub use | 291, 303, 304 | F10-F19 |
| schizophrenia spectrum | 295, 297, 298 | F20-F29 |
| schizophrenia | 295 | F20 |
| *Any mood* | 296 | F30-F39 |
| bipolar disorder | 296.19, 296.39, 298.19 | F30-31 |
| depression | 296.09, 296.29, 298.09, 300.49 | F32-33 |
| neurotic/stress disorder | 300, 305 | F40-F48 |
| OCD | 300.3 | F42.0, F42.1, F42.2 |

| | | |
|---|---|---|
| **behavioral synd-physiol** | **306.49, 306.50, 305.58, 306.59** | **F50-F59** |
| **anorexia nervosa** | **306.5** | **F50.0** |
| **adult personality disorder** | **301, 302** | **F60-F69** |
| **intellectual disability** | **310-315** | **F70-F79** |
| **psych dev dis - not ASD** | **306.10. 306.11. 306.12, 306.18, 306.19** | **F80-F89 (not F84.0, F84.1, F84.5, F84.8, F84.9)** |
| **behav dis-child onset** | **308, 306.09, 306.29, 306.79, 306.89** | **F90-F98** |
| **ADHD** | **NA** | **F90, F98.8** |
| **tic disorder** | **306.2** | **95.1, 95.2** |
| **mental-unspecified** | **NA** | **F99** |
| **<u>Neurologic disorders</u>** | | |
| ***Any neurologic*** | **320-358** | **G00-G99** |
| **inflammatory of CNS** | **320-324** | **G00-G09** |
| **systemic atrophies** | **331.09, 332, 348.09, 348.20, 348.29, 348.99** | **G10-G14** |
| **extrapyramidal** | **342** | **G20-G26** |
| **other degenerative** | **NA** | **G30-G32** |
| **demyelinating of CNS** | **340, 341.01** | **G35-G37** |
| **episodic** | **345, 346, 347.00, 347.01, 347.09** | **G40-G47** |
| **epilepsy** | **345** | **G40-G41** |
| **nerve disorder** | **350-358** | **G50-G59** |
| **polyneuropath** | **354** | **G60-G64** |
| **myoneural junction** | **330, 733.09** | **G70-G73** |
| **cerebral palsy** | **343** | **G80-G83** |

| | | |
|---|---|---|
| other neurologic | 347.93, 347.94, 347.95, 349.00, 349.01, 349.09 | G90-G99 |
| <u>Cardiometabolic disorders</u> | | |
| Type 2 diabetes | 250 | E11, O24.1 |
| gestational diabetes | 761.1 | O24.4 |
| *Any diabetes* | 761.1, 250, 249 | E10, E11, E12, E13, E14, G59.0, G63.2, H28.0, H36.0, M14.2, N08.3, O24.0, O24.1, O24.2, O24.3, O24.4, O24.9 |
| obesity | 277, 277.99,278 | E66, O99.21; Z68.2, Z68.3, Z68.4 |
| diabetes outside preg | 249, 250 | E10, E11, E12, E13, E14, G59.0, G63.2, H28.0, H36.0, M14.2, N08.3, O24.0, O24.1, O24.2, O24.3, O24.9 |
| preeclampsia/eclampsia | 637.0, 637.1, 762.1, 762.2, 637.9 | O11, O14, O15 |
| hypertension in preg | 760.2, 637.0, 637.1, 762.1, 762.2, 637.9 | O10-O16, excluding O12 |
| *Any hypertension* | 400-404, 760.2, 637.0, 637.1, 637.9, 762.1, 762.2 | I10-I15, O10-O16, excluding O12 |
| hypertension outside preg | 400-404 | I10-I15 |
| Type 1 diabetes | 249 | E10, O24.0 |
| <u>Birth defects</u> | | |
| *Any birth defect* | 740-759 | Q00.x-Q99.x (excluding undescended testicle Q53 and congenital deformities of hip Q65). DO include these codes in birth defects as a whole : ICD10 D18.02, D18.03, D27, E25.0, E34.5, G52.7, K43.9; ICD8 220.0, 553.2, 553.3, 553.8, 553.9 |

| | | |
|---|---|---|
| CNS | 740-743 | Q00-Q07 |
| eye | 744 | Q10-Q15 |
| ear | 745 | Q16-Q18 |
| heart | 746-747 | Q20-Q28 |
| respiratory | 748 | Q30-Q34 |
| lip | 749 | Q35-Q37 |
| digestive system | 750-751 | Q38-Q45 |
| genital | 752 | Q50-Q56 |
| urinary tract | 753 | Q60-Q64 |
| musculoskeletal | 754-756 | Q65-Q79 |
| skin | 757 | Q80-Q84 |
| other/chromosomal | 758-759 | Q85-99 |
| chromosomal/gene dis- ASD specific | 759.83, 759.30, 759.51 | D82.1, G71.0, Q85.0, Q85.1, Q87.1, Q87.8, Q90, Q93.5, Q93.8, Q98.0-98.4, Q99.2 |
| <u>Autoimmune disorders</u> | | |
| Type 1 diabetes | 249 | E10; O24.0 |
| thyrotoxicosis | 242 | E05.0 |
| thyroiditis | 245.03 | E06.3 |
| primay adrenocortical | 255.1 | E27.1 |
| rheumatoid arthritis | 712.19, 712.39, 712.59 | M05, M06 |
| juvenile arthritis | 712.09 | M08 |
| dermatopolymyositis | 716 | M33 |
| polymyalgia | 446.3 | M31.5, M31.6, M35.3 |
| scleroderma | 734 | M34 (except M34.2) |
| lupus erythema | 734.19 | M32.1, M32.8, M32.9 |
| sjogren | 734.9 | M35.0 |
| ankylos spondil | 712.49 | M45.9 |
| Wegner granulomatosis | 446.29 | M31.3 |
| celiac | 269.00 | K90.0 |
| crohn | 563.01 | K50 |

| | | |
|---|---|---|
| ulcerative colitis | 563.19 | K51 |
| pernicious anem | 281.0 | D51.0 |
| hemolytic anem | 283.90. 283.91 | D59.1 |
| purpura | 446.49 | D69.3 |
| multiple sclerosis | 340 | G35 |
| guillain-bar | 354 | G61.0 |
| Myasthen grav | 733.09 | G70.0 |
| pemphigus | 694 | L10 (except L10.5) |
| psoriasis | 696.09, 696.10, 696.19 | L40 (except L40.4) |
| alopecia areata | 704 | L63 |
| vitiligo | 709.01 | L80.9 |
| *any endocrine* | 249, 242, 245.03, 255.1 | E10; O24.0, E05.0, E06.3, E27.1 |
| *any connective* | 712.19, 712.39, 712.59, 712.09, 716, 446.3, 734, 734.19, 734.9, 712.49, 446.29 | M05, M06, M08, M33, M31.5, M31.6, M35.3, M34 (except M34.2), M32.1, M32.8, M32.9, M35.0, M45.9, M31.3 |
| *any gastrointestinal* | 269.00, 563.01, 563.19 | K90.0, K50, K51 |
| *any blood* | 281.0, 283.90, 283.91, 446.49 | D51.0, D59.1, D69.3 |
| *any nervous* | 340, 354, 733.09 | G35, G61.0, G70.0 |
| *any skin* | 694, 696.09, 696.10, 696.19, 704.00, 709.01 | L10 (except L10.5), L40 (except L40.4), L63, L80.9 |
| *any autoimmune* | 249, 242, 245.03, 255.1, 712.19, 712.39, 712.59, 712.09, 716, 446.3, 734, 734.19, 734.9, 712.49, 446.29, 269.00, 563.01, 563.19, 281.0, 283.90, 283.91, 446.49, 340, 354, 733.09, 694, 696.09, 696.10, 696.19, 704.00, 709.01 | E10; O24.0, E05.0, E06.3, E27.1, M05, M06, M08, M33, M31.5, M31.6, M35.3, M34 (except M34.2), M32.1, M32.8, M32.9, M35.0, M45.9, M31.3, K90.0, K50, K51, D51.0, D59.1, D69.3, G35, G61.0, G70.0, L10 (except L10.5), L40 (except L40.4), L63, L80.9 |

| | | |
|---|---|---|
| **Asthma** | 493.00, 493.01, 493.02, 493.08, 493.09 | J45, J46 |
| **Allergies** | 493.02, 507, 691.00, 999.49, 708.09 | J45.0, J30, L20, T78.0, T78.1, T78.2, T78.3, T78.4, H10.1 |

**eFigure 1. Selection of study cohort**

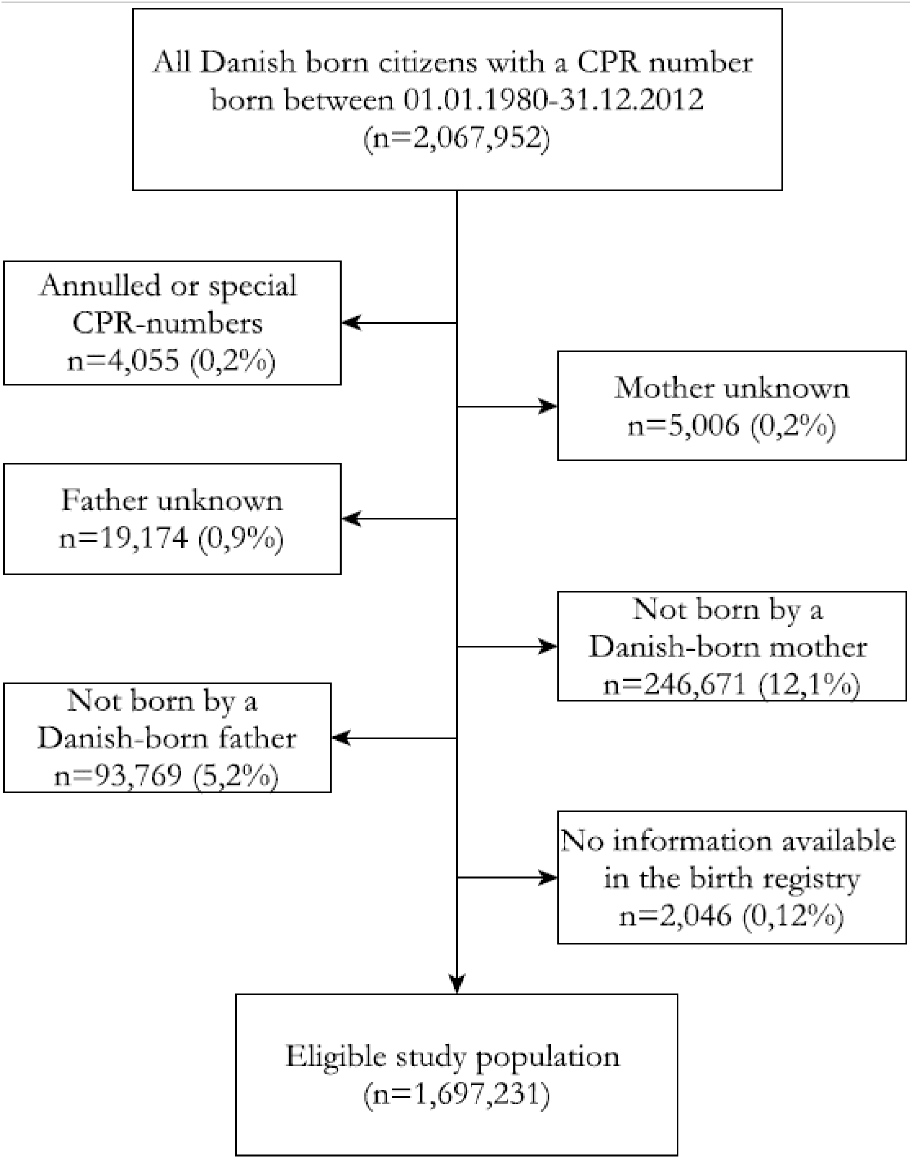

**eFigure 2. Overview of family member types and diagnosis categories for analysis and risks to be estimated**

**A. Adjusted risk estimates for ASD in the index person when a family member (type1, …., type 20), has a reported diagnosis of (dx 1, ,….., dx 90).**

| | 20 family member types of different degrees of relatedness, sex and side of the family | | | 90 diagnosis categories[a] | |
|---|---|---|---|---|---|
| **Risk for:** | **1st degree relative (50% shared genes with index person)** | **2nd degree relative (25% shared genes with index person)** | **3rd degree relative (12.5% shared genes with index person)** | **Major diagnosis group** | **Number of specific diagnosis categories per major group** |
| ASD overall | Mother | Maternal grandmother | Maternal cousin, female[b] | **Mental** | 20 diagnoses |
| ASD in females | Father | Maternal grandfther | Maternal cousin, male[b] | **Neurologic** | 13 diagnoses |
| ASD in males | Full sibling, female | Maternal aunt[b] | Paternal cousin, female[b] | **Cardiometabolic** | 10 diagnoses |
| ASD with ID | Full sibling, male | Maternal uncle[b] | Paternal cousin, male[b] | **Birth defect** | 14 diagnoses |
| | | Maternal half sibling, female[b] | | **Autoimmune** | 31 diagnoses |
| | | Maternal half sibling, male[b] | | **Asthma/allergies** | 2 diagnoses |
| | | Paternal grandmother[b] | | | |
| | | Paternal grandfather[b] | | | |
| | | Paternal aunt[b] | | | |
| | | Paternal uncle | | | |
| | | Paternal half sibling, female[b] | | | |
| | | Paternal half sibling, male[b] | | | |

**B. Adjusted risk estimate for a co-occuring condition (dx 1,….. dx 89) in a person with ASD**

| **Risk for co-ocurring diagnoses in persons with ASD, by ASD phenotype and sex** | 89 co-occuring diagnosis categoriess[a] | |
|---|---|---|
| | **Major group** | **Number of specific diagnosis categories per major group** |
| Females with ASD | **Mental** | 19 diagnoses |
| Males with ASD | **Neurologic** | 13 diagnoses |
| Females with ASD and ID | **Cardiometabolic** | 10 diagnoses |
| Males with ASD and ID | **Birth defect** | 14 diagnoses |
| | **Autoimmune** | 31 diagnoses |
| | **Asthma/allergies** | 2 diagnoses |

[a]See eTable 3 for specific ICD8 and ICD10 codes used to identify each specific diagnosis. For index persons, siblings, cousins; diagnoses reported before the end of study followup were included. For all other family members types: diagnoses reported before birth of the index child were included.

[b]Earliest reported diagnosis used if 2 or more family members of the same type had the diagnosis

**eFigure 3. Percent of study cohort members with linkages to siblings, grandparents, aunts, uncles and cousins by birth year**

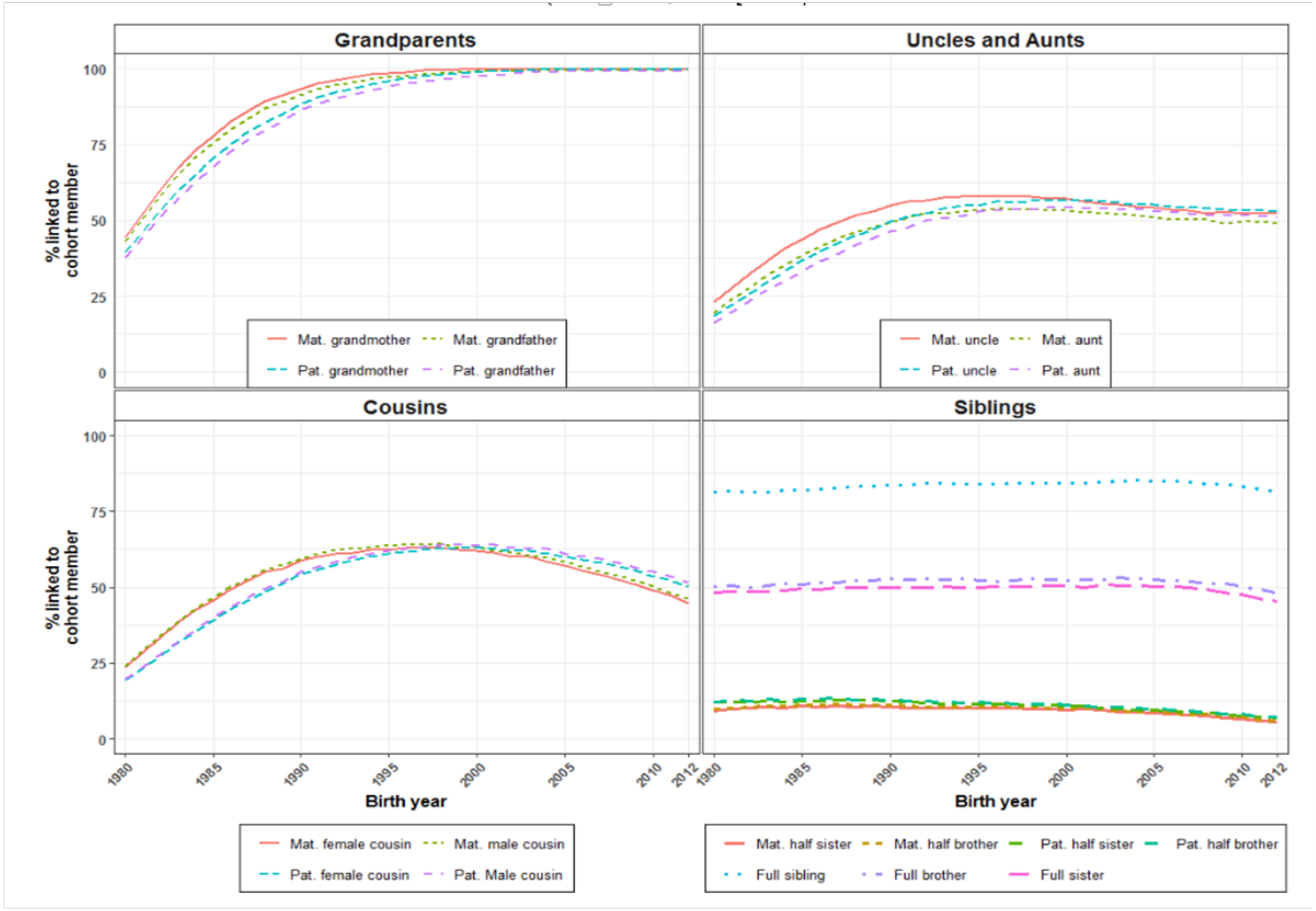

**eFigure 4. Risk for ASD (aHR[a]) for ASD from family morbidity by diagnosis and family member type, statistically significant estimates only (aHR confidence interval excludes 1.0)**

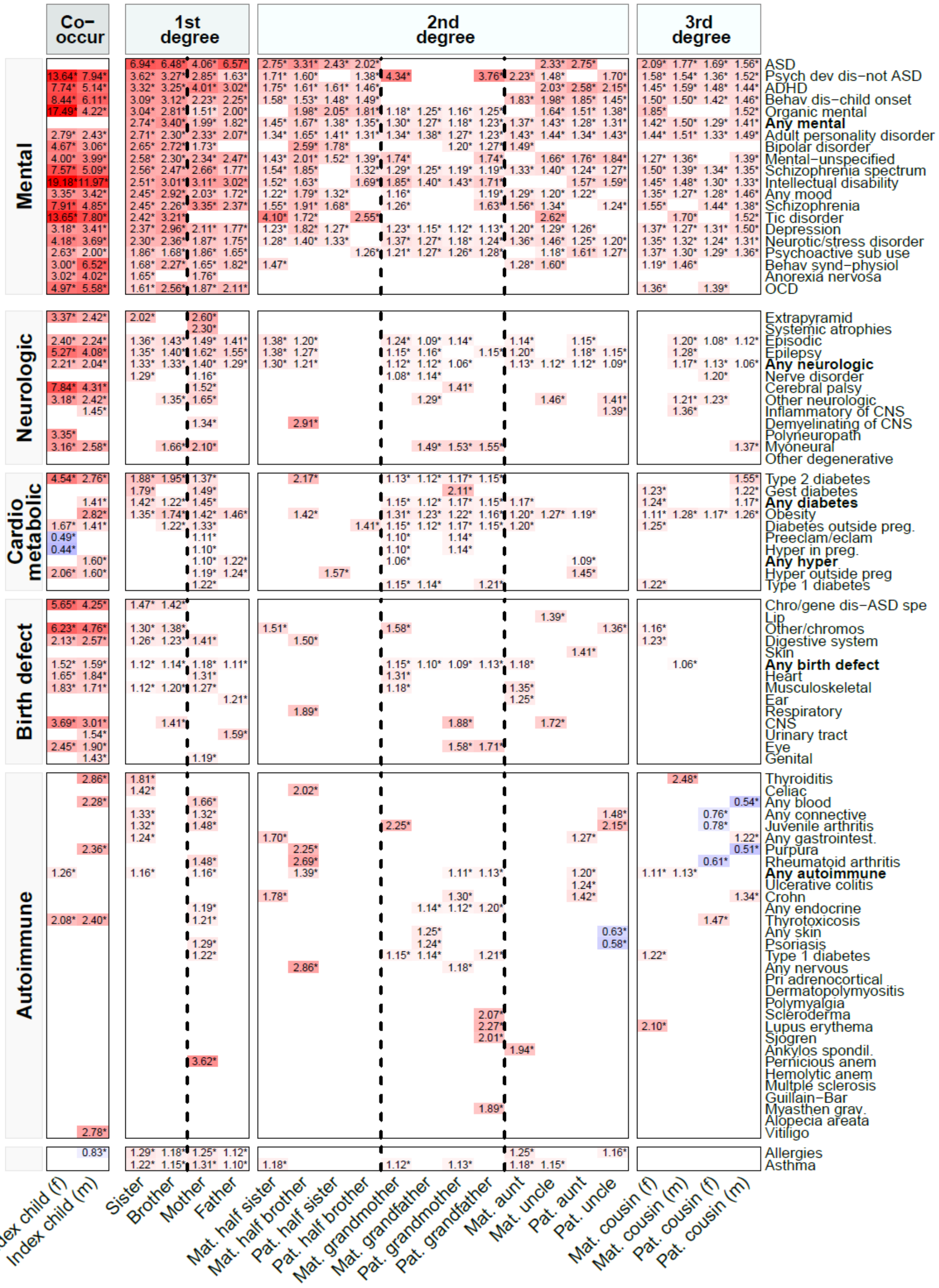

<u>**eFigure 4 legend**</u>

**Hazard Ratio: the value in a cell of the heat map corresponds to the adjusted Hazard Ratio for ASD estimated from that family member type-morbidity combination. Co-occurring condition: the two co-occurring condition columns correspond to risk of co-occurrence of a specific diagnosis in autistic females and males. 1st degree, 2nd degree, 3rd degree: each column corresponds to risk for ASD from a specific family member type (such as a full sister) with a specific diagnosis by degree of relatedness. 1st degree: 50% shared genes with index person. 2nd degree: 25% shared genes with index person. 3rd degree: 12.5% shared genes with index person. Note on blank cells: a blank cell reflects 1) a sample size of ASD persons of <5 for the specific family member type-diagnosis combination (or <5 ASD persons with the specific co-occurring diagnosis) or 2) an aHR with 95% confidence interval that includes 1.0. Row order: row order the same as in Figure 1.**

**[a]Hazard Ratio (aHR) adjusted for sex, birth weight, gestational age, parental ages at birth and labeled significant if 95% confidence interval excluded 1.0. Each aHR estimated using separate baseline ASD --diagnostic rates per birth year. aHRs from full/half siblings, aunts, uncles or cousins also included the number of family members of the relevant type as a covariate.**

### eFigure 19. Risk for ASD (aHR[a]) in males from family morbidity by diagnosis and family member type

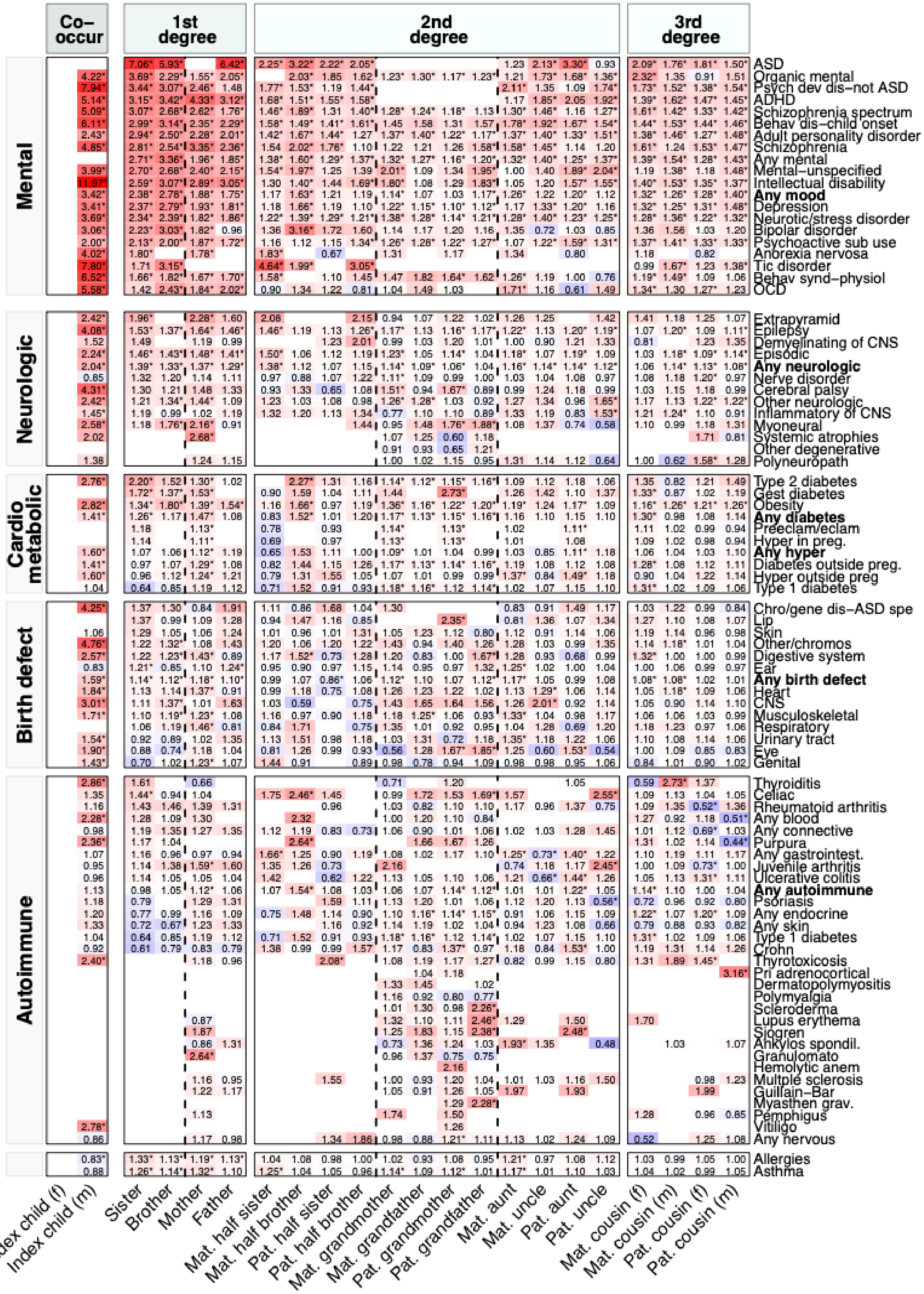

**eFigure 19 legend**

**Hazard Ratio: the value in a cell of the heat map corresponds to the adjusted Hazard Ratio for ASD in males estimated from that family member type-morbidity combination. Co-occurring condition: the male Co-occurring condition column corresponds to risk of co-occurrence of a specific diagnosis in autistic males. 1st degree, 2nd degree, 3rd degree: each column corresponds to risk for ASD in males from a specific family member type (such as a full sister) with a specific diagnosis by degree of relatedness. 1st degree: 50% shared genes with index person. 2nd degree: 25% shared genes with index person. 3rd degree: 12.5% shared genes with index person. Note on blank cells: a blank cell reflects a sample size of male autistic persons of <5 for the specific family member type-diagnosis combination (or <5 male autistic persons with the specific co-occurring diagnosis). Note on row order: Within each major disorder group, the rows are ordered by diagnosis with the largest aHR (top row) to the smallest aHR (bottom row) from full sisters.**

**[a]Hazard Ratio (aHR) adjusted for birth weight, gestational age, parental ages at birth. Each aHR estimated using separate baseline ASD diagnostic rates per birth year. aHRs from full/half siblings, aunts, uncles or cousins also included the number of family members of the relevant type as a covariate.**

### eFigure 20. Risk for ASD (aHR[a]) in females from family morbidity by diagnosis and family member type

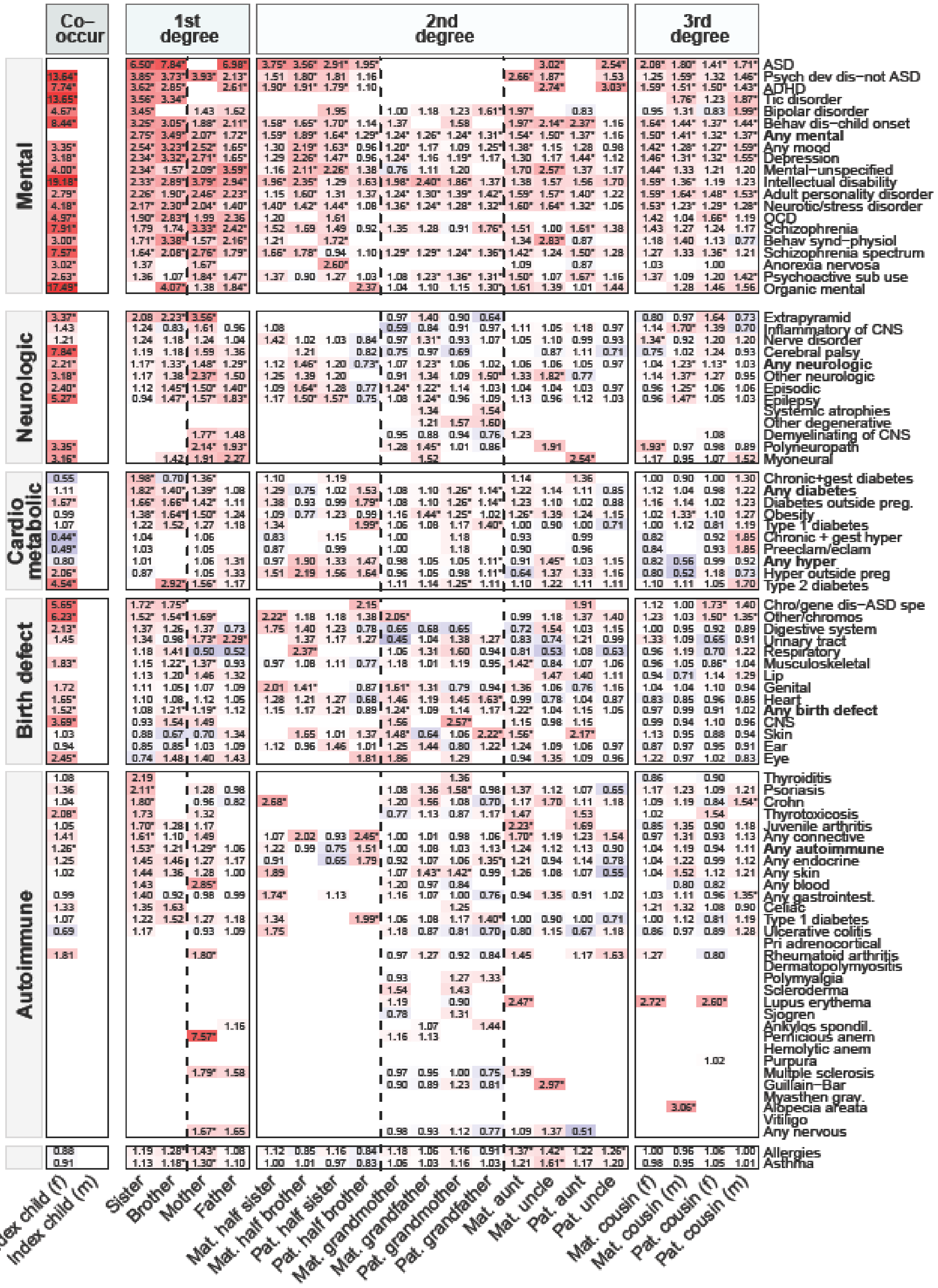

**eFigure 20 legend**

**Hazard Ratio: the value in a cell of the heat map corresponds to the adjusted Hazard Ratio for ASD in females estimated from that family member type-**

**morbidity combination. Co-occurring condition: the female Co-occurring condition column corresponds to risk of co-occurrence of a specific diagnosis in autistic females. 1st degree, 2nd degree, 3rd degree: each column corresponds to risk for ASD in females from a specific family member type (such as a full sister) with a specific diagnosis by degree of relatedness. 1st degree: 50% shared genes with index person. 2nd degree: 25% shared genes with index person. 3rd degree: 12.5% shared genes with index person. Note on blank cells: a blank cell reflects a sample size of female autistic persons of <5 for the specific family member type-diagnosis combination (or <5 female autistic persons with the specific co-occurring diagnosis). Note on row order: Within each major disorder group, the rows are ordered by diagnosis with the largest aHR (top row) to the smallest aHR (bottom row) from full sisters.**

**[a]Hazard Ratio (aHR) adjusted for birth weight, gestational age, parental ages at birth. Each aHR estimated using separate baseline ASD diagnostic rates per birth year. aHRs from full/half siblings, aunts, uncles or cousins also included the number of family members of the relevant type as a covariate.**

**eFigure 56. Risk for ASD (aHR[a]) with intellectual disability (ID) from family morbidity by diagnosis and family member type**

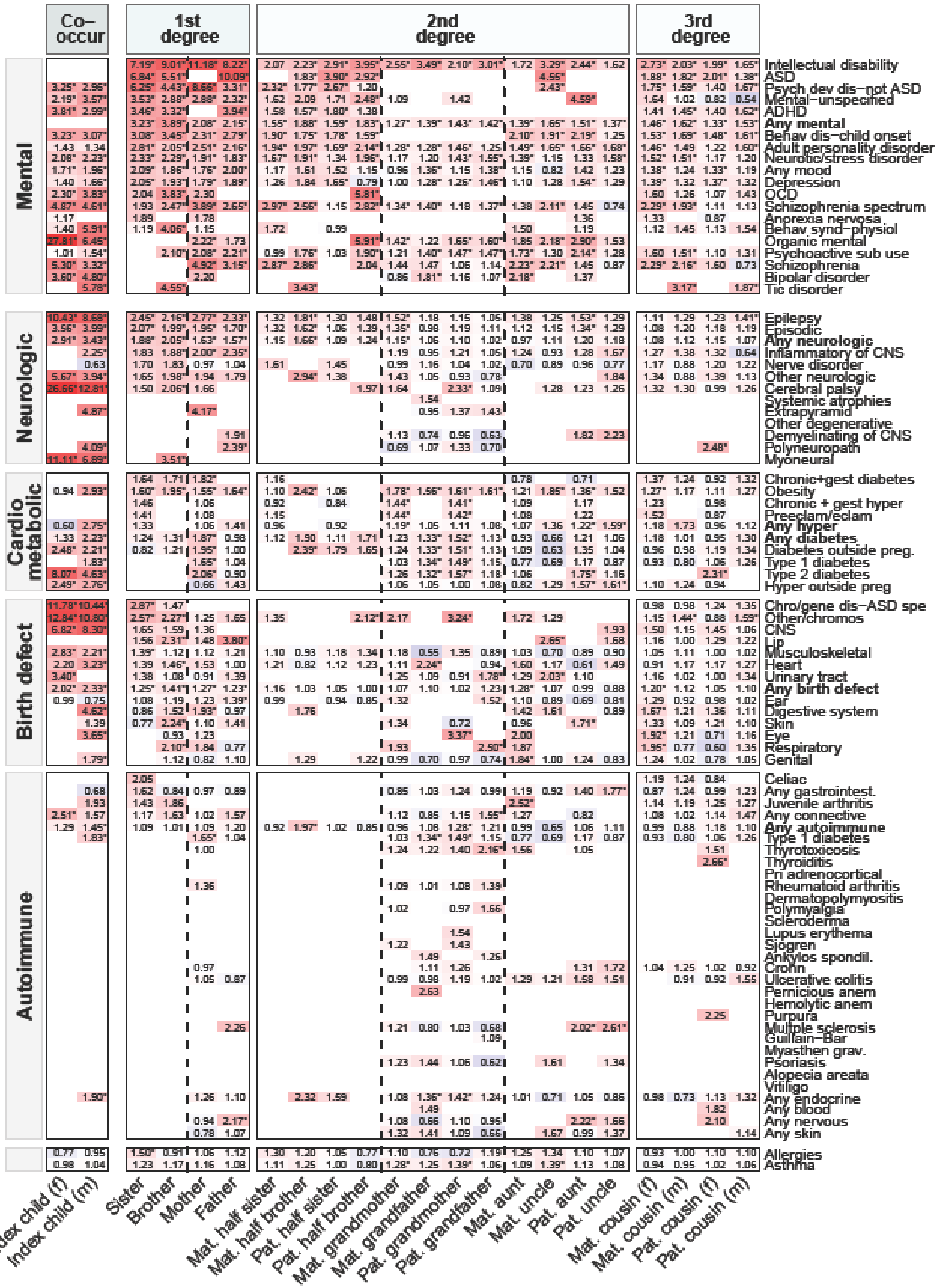

### eFigure 56 legend

**Hazard Ratio: the value in a cell of the heat map corresponds to the adjusted Hazard Ratio for ASD with ID estimated from that family member type-**

**morbidity combination. Co-occurring condition: the two Co-occurring condition columns correspond to risk of co-occurrence of a specific diagnosis in autistic females and males with ID. 1st degree, 2nd degree, 3rd degree: each column corresponds to risk for ASD with ID from a specific family member type (such as a full sister) with a specific diagnosis by degree of relatedness. 1st degree: 50% shared genes with index person. 2nd degree: 25% shared genes with index person. 3rd degree: 12.5% shared genes with index person. Note on blank cells: a blank cell reflects a sample size of ASD with ID persons of <5 for the specific family member type-diagnosis combination (or <5 ASD with ID persons with the specific co-occurring diagnosis). Note on row order: Within each major disorder group, the rows are ordered by diagnosis with the largest aHR (top row) to the smallest aHR (bottom row) from full sisters.**

**[a]Hazard Ratio (aHR) adjusted for sex, birth weight, gestational age, parental ages at birth; each aHR estimated using separate baseline ASD diagnostic rates per birth year. aHRs from full/half siblings, aunts, uncles or cousins also included the number of family members of the relevant type as a covariate.**
